# Higher Limbic and Basal Ganglia volumes in surviving COVID-negative patients and the relations to fatigue

**DOI:** 10.1101/2021.11.23.21266761

**Authors:** Rakibul Hafiz, Tapan Kumar Gandhi, Sapna Mishra, Alok Prasad, Vidur Mahajan, Xin Di, Benjamin H. Natelson, Bharat B. Biswal

## Abstract

**Background:** Among systemic abnormalities caused by the novel coronavirus, little is known about the critical attack on the central nervous system (CNS). Few studies have shown cerebrovascular pathologies that indicate CNS involvement in acute patients. However, replication studies are necessary to verify if these effects persist in COVID-19 survivors more conclusively. Furthermore, recent studies indicate fatigue is highly prevalent among ‘long-COVID’ patients. How morphometry in each group relate to work-related fatigue need to be investigated.

**Method:** COVID survivors were MRI scanned two weeks after hospital discharge. We hypothesized, these survivors will demonstrate altered gray matter volume (GMV) and experience higher fatigue levels when compared to healthy controls, leading to stronger correlation of GMV with fatigue. Voxel-based morphometry was performed on T1-weighted MRI images between 46 survivors and 30 controls. Unpaired two-sample t-test and multiple linear regression were performed to observe group differences and correlation of fatigue with GMV.

**Results:** The COVID group experienced significantly higher fatigue levels and GMV of this group was significantly higher within the *Limbic System* and *Basal Ganglia* when compared to healthy controls. Moreover, while a significant positive correlation was observed across the whole group between GMV and self-reported fatigue, COVID subjects showed stronger effects within the *Posterior Cingulate, Precuneus* and *Superior Parietal Lobule*.

**Conclusion:** Brain regions with GMV alterations in our analysis align with both single case acute patient reports and current group level neuroimaging findings. We also newly report a stronger positive correlation of GMV with fatigue among COVID survivors within brain regions associated with fatigue, indicating a link between structural abnormality and brain function in this cohort.

## Introduction

SARS-CoV-2 (Severe Acute Respiratory Syndrome Coronavirus 2) is a highly contagious novel coronavirus that started a global pandemic causing more than 400 million confirmed cases of COVID-19 and nearly 6 million deaths world-wide[1]. Mass vaccinations have mitigated cases in USA, but infections continue to rise – especially in India. The literature shows evidence of structural brain abnormalities but exactly where and how long these effects persist in acute patients during recovery, remains unclear.

Attack on the central nervous system (CNS) in acutely ill COVID-19 patients can cause a broad range of pathology including ischemic strokes, encephalitis [2], inflammatory vascular pathologies in cerebral vessels [3, 4], and microhemorrhages [5] among many others. Ischemic strokes (27%), encephalitis (13%), confusion (53%), impaired consciousness (39%) along with agitation (31%) and headaches (16%) were also reported from patients across multiple sites (11 hospitals, n = 64) [2]. Moreover, brain lesions and hyperintensities were observed from fluid-attenuated inversion recovery (FLAIR) imaging in multiple brain regions [6] along with autoimmune and hemorrhagic encephalitis [7]. But these were mostly case studies, and the current literature needs to move from individual cases to conclusive group level estimates delineating structural brain alterations between COVID-19 patients and healthy controls (HCs).

A few recent neuroimaging studies have emerged to address this gap with moderate [8, 9] to large sample sizes [10], including follow-up [11, 12] and longitudinal designs [10]. Lu et al., 2020 [11] had reported neurological symptoms in over 68% (41/60) of hospitalized patients that persisted after a 3 month follow up (55%). They had also performed structural MRI and showed significantly higher gray matter volume (GMV) in several regions of interest (ROIs) – Rolandic operculum, bilateral olfactory, insular, and hippocampal regions, as well as in the right cingulate gyrus and left Heschl’s gyrus [11]. An alarming number of survivors are now undergoing a sequela of symptoms [13-15] which converge to the brain as the responsible organ. Therefore, changes in brain structure could correlate to the severity of these symptoms. In regard to that, another follow-up study [12] assessed structural and functional changes in COVID-19 patients in two consecutive time points - 3 and 6 months and evaluated their relation to post-traumatic stress symptoms (PTSS). They showed increased GMV in bilateral hippocampus and amygdala, which also correlated negatively with self-reported Posttraumatic Stress Disorder Checklist for DSM-5 (PCL-5) scores in COVID subjects. This could also relate to the interval after hospital discharge and the severity of symptoms can alter with time. For example, Tu et al., also show that the PCL-5 scores from Session1 (3 months) correlated with the time after discharge and the total PCL-5 scores from these survivors increased by ∼20% at Session2 (6 months).

It is possible that the COVID and control groups do not demonstrate any global GMV changes, rather, GMV could be modulated locally due to fever and hypoxemic conditions. For instance, a recent study used source-based morphometry (SBM), a multivariate alternate to VBM, on Computed Tomography (CT) scans to show that the fronto-temporal network is more susceptible to fever and reduced oxygen levels in COVID patients, despite no global difference in GMV [8]. Furthermore, the modified Rankin Scale (mRS), a clinical disability score, significantly correlated with lower GMV in the frontal gyrus both during discharge and after a 6 month-follow-up. Interestingly, COVID survivors can also develop neurological symptoms during recovery, despite no such manifestations in the acute stage. For example, a recent study reported reduced cortical thickness in the left insula, hippocampus, and superior temporal gyrus, after a 3 month-follow-up MRI scan in two 2 sub-types (mild and severe), who had no signs of neurological manifestations during the acute stage [9],

Another important question is how these neurological changes develop in individuals before and after infection with COVID-19? A recent longitudinal study [10] used a large pool of patients (N=785, n_COVID_ = 401) from the UK Biobank COVID-19 reimaging study, to show reduced GM thickness and contrast in the orbitofrontal and parahippocampal gyrus, as well as, in insula, amygdala and the anterior cingulate cortex. In addition, they report increased tissue damage in brain regions functionally associated with the piriform cortex and the olfactory system, as well as higher volumes in CSF. Therefore, the literature shows mixed evidence of increased GMV and contrarily, reduced GM thickness from cross-sectional, follow-up and longitudinal designs, nevertheless, in quite consistent anatomical locations. Our goal was to first assess, if the participants from our study, scanned after a much shorter interval from hospital discharge (2 weeks), demonstrated altered GMV in regions that are consistent with both acute stage single case reports and more recent group level neuroimaging reports from lengthy recovering (3 to 6 months) patients. Moreover, since fatigue is the highest reported symptom from surviving patients [13-15], we wanted to ask, if self-reported fatigue (during work) independently correlated to voxel-wise GMV in regions, known to be functionally associated with fatigue.

In this study, we try to address this by recruiting a group of patients, hospitalized due to a positive PCR test for COVID-19. We imaged them two weeks after hospital discharge after converting to be PCR negative. One expectation is that changes in brain tissue structure in COVID survivors can still cause changes in compartmental volumes that remain shortly after hospital discharge. Specifically, we hypothesized that these surviving COVID-negative patients would demonstrate GMV differences with the HC group and show significant correlation of altered GMV with self-reported fatigue scores. T1-weighted MRI images are sensitive to such changes and can be used to estimate GMV differences between two groups using voxel-based morphometry (VBM) [16].

## Materials and Methods

### Participants

47 COVID-negative patients and 35 HCs were recruited by the Indian Institute of Technology (IIT), Delhi, India where they were imaged following all Institutional Review Board (IRB) guidelines. Please note, these subjects were recruited from a much larger pool of patients. The patients were initially classified based on illness severity data derived from a database of 2,538 COVID patients admitted to the Metro Hospital in Delhi from May to December 2020. 24% of this sample did not require O_2_, 40% required O_2_, 22% required Continuous Positive Airway Pressure (CPAP); and 14% were intubated. This 14% of intubated patients were excluded from the recruitment process in the current study. Patients were studied two weeks after discharge, after becoming PCR negative [333 who needed CPAP to raise O_2_ levels; 333 who needed nasal O_2_ to raise O_2_ levels; and 334 who were admitted but did not need supplemental O_2_]. The sample of 47 COVID subjects constituting the COVID group in this study were collected from this cohort (those who agreed to participate in this ongoing study so far). Of these 47, 36.17% (17/47) patients were reported to be ‘mild’, 8.51% (4/47) to be ‘moderate’ and 36.17% (17/47) were between ‘moderate’ to ‘severe’. Information from the rest of the 19.15% (9/47) was not provided by the hospital because those patients did not give consent to sharing their medical symptoms. Among the ‘moderate’ to ‘severe’ patients, 58.82% (10/17) were given Remdesivir and 29.41% (5/17) required additional oxygenation. One patient (1/17) was given a mix of Dexamethasone, Ceftriaxone and Clexane injections and another (1/17) was put in an Antibiotic and Steroid regime for 4 weeks (Progressively reduced). One other patient (1/17) was given Actemera 2 times, who was also administered Bilevel Positive Air Pressure (BiPap) for 4-5 days. On average these 47 patients stayed in the hospital for approximately 11 ± 3.30[SD] days. Before data analysis, six subjects (1 COVID and 5 HC) were removed during quality control assessment. Effectively, T1-weighted images from 46 (31 males) COVID and 30 (23 males) HCs were included in the study with mean age 33.5 years ± 9.74[SD] years (HC) and 34.63 years ±11.54[SD] years (COVID). Please see *Table 1* for more details on demographics.

**Table 1:** Group level statistics on participant demographics and global VBM metrics of each tissue type in HC and COVID group. The first three rows represents the test results from participant demographics including age, sex and fatigue. The last four rows represents the global VBM metrics of compartmental and total volumes. Keys: GMV = Gray Matter Volume, WMV = White Matter Volume, CSFV = Cerebrospinal Fluid Volume, TIV = Total Intracranial Volume, *p = p-value, t = two-sample t-test statistic, χ*^2^*= Chi-Squared statistic, T = Wilcoxon Rank Sum test score*, ml = milliliter, M = Male, F = Female.

| Measures | *p* | *stat* | HC, mean (SD) | COVID, mean (SD) |
| --- | --- | --- | --- | --- |
| Age (years) | 0.66 | -0.44 (*t*) | 33.50 (9.74) | 34.63 (11.54) |
| Sex | 0.38 | 0.76 ($\chi^2$) | 23M, 7F | 31M, 15F |
| Fatigue | 2.86e-07 | 1093 (*T*) | 0.61 (0.78) | 2.67 (1.27) |
| GMV (ml) | 0.57 | -0.57 (*t*) | 629.59 (44.17) | 638.30 (76.39) |
| WMV (ml) | 0.20 | -1.30 (*t*) | 394.44 (42.51) | 407.64 (43.93) |
| CSFV (ml) | 0.20 | -1.29 (*t*) | 246.49 (59.72) | 263.48 (54.02) |
| TIV (ml) | 0.20 | -1.30 (*t*) | 1270.52 (106.92) | 1309.42 (139.03) |

### Clinical Assessment

The most commonly reported symptoms from the participants during hospitalization were - fever, cough, body ache, chills, difficulty breathing, bowel irritation, nausea, loss of sense of smell and loss of consciousness. We also assessed if they were having any ongoing/new symptoms from day of discharge to the day of scan – fatigue, anxiety, lack of attention, body ache, headache, memory loss, delayed recovery of sense of taste and/or smell, bowel irritation and interestingly, hair loss were commonly reported. Particularly, since we were interested in fatigue related correlates of GMV, a subset of each group (n_COVID_ = 33, n_HC_ = 18), successfully reported their fatigue levels on a scale of 0 to 5, with 0 representing no fatigue and 5 representing the highest fatigue levels observed during work. The average fatigue score in the sub-set of COVID participants was 2.67/5 ± 1.27 [SD] and that of the HC group was 0.61 ± 0.78[SD].

### Imaging

High-resolution structural images were acquired using a 3T GE scanner with a 32-channel head coil in 3D imaging mode with a fast BRAVO sequence. The imaging parameters were TI = 450 ms; 244 × 200 matrix; Flip angle = 12 and FOV = 256 mm, number of slices = 152 (sagittal), slice thickness = 1.00 mm and spatial resolution of 1.0 mm × 1.0 mm × 1.0 mm.

### Data Pre-Processing

We performed pre-processing using SPM12 (http://www.fil.ion.ucl.ac.uk/spm/) within the MATLAB environment (Mathworks Inc, Massachusetts, USA). All anatomical images were visually inspected for artifacts, re-centered and reoriented to the anterior-posterior commissure (ac-pc) line. Each brain compartment was segmented into specific tissue classes mainly - gray matter (GM), white matter (WM), and cerebro-spinal fluid (CSF). A study-specific template was first generated using the fast diffeomorphic image registration algorithm (DARTEL) [17] which is representative of the average across all the participants included in the study [18, 19]. Subject level maps were non-linearly warped to this reference template for relatively higher specificity and accuracy [19]. Finally, each map was normalized to the Montreal Neurological Institute (MNI) space using affine transformation and resampled to an isotropic voxel dimension of 1.5 mm × 1.5 mm × 1.5 mm. Modulated images were obtained for each subject, which account for contractions and expansions from non-linear spatial transformations. The normalized modulated images were then spatially smoothed with a gaussian kernel of 8 mm. VBM is a volumetric computational method that can quantify voxel-wise changes in tissue volume in the gray matter (GM)[16]. It is a useful method to report group level differences in tissue volume between patients and healthy controls (HCs), using T1-weighted anatomical images. VBM can be estimated from gray matter probability maps obtained from the segmentation stage. Each value in a tissue specific probability map represents the likelihood of the voxel belonging to a brain compartment and tissue volumes can estimated by summing over the product of each voxel’s dimension and the corresponding probabilities [20]. Total Intracranial Volume (TIV) is the sum of volumes from each major compartment in the brain – GM, WM, and CSF, with TIV = GMV + WMV + CSFV; where GMV = Gray Matter Volume, WMV = White Matter Volume and CSFV = Cerebro-Spinal Fluid Volume.

These quantities were used to assess central tendency measures in each group. To visualize the probability distributions and group average compartmental and total brain volumes, we customized and adopted a script in RStudio [21] to generate a ‘raincloud’ figure, as depicted in a recent publication [22].

### Statistical Analysis

To assess differences in participant demographics, we performed two sample t-test on age, GMV, WMV, CSFV and TIV and chi-squared test for sex differences between the two groups. Since the fatigue scores deviated from normality (Shapiro Wilk: p < 0.05), we used Wilcoxon’s ranksum test to assess group difference between HCs and COVID subjects. To determine group level differences in GMV, we performed a two-sample t-test using the smoothed, modulated, and normalized GM tissue maps from the two groups. TIV of each subject was group-mean centered and added as a covariate along with age and sex to account for confounding effects. An implicit mask with absolute threshold of 20% above the group mean was set to exclude unwanted voxel quantities from the smoothed GM maps. Regions with significant difference in GMV between the two groups were identified by first applying a height threshold of *p*_*unc*_ *< 0*.*001* and then *family wise error (FWE)* corrected at *p*_*FWE*_ *< 0*.*05*, for multiple comparisons. Cluster-based thresholding can be a problem when using VBM since residuals tend to vary in spatial smoothness. Therefore, we used a non-stationary cluster-based correction available in SPM to evaluate significant results.

To evaluate the relationship between GMV and fatigue in the sub-set of COVID participants, we performed a multiple linear regression analysis, with voxel-wise GMV as the response variable and the fatigue score as the covariate of interest, while, age, sex and TIV were included as covariates of no interest. Regions with significant correlation between GMV and fatigue score were identified using cluster-based thresholding at height threshold *p*_*unc*_ *< 0*.*001* and *FWE* corrected at *p*_*FWE*_ *< 0*.*05*, for multiple comparisons. In order to visualize any significant linear relationship between the two variables, the average GMV within the significant cluster was obtained from each subject. These average GMV values were then linearly regressed against the fatigue scores and visualized within a scatter plot and a line of best fit with 95% confidence interval. Age, sex and TIV of each participant were regressed out during the linear regression step. The mean GMV across the participants was then added back to the residuals and the correlation with fatigue scores was computed. The correlation analysis and the graphical plotting was done using ‘inhouse’ scripts prepared in RStudio [21].

## Results

We assessed group level differences in demographics (age, sex and fatigue), as well as in global VBM metrics including GMV, WMV, CSFV and TIV. We observed no significant differences in age and sex between the two groups *(p > 0*.*05)*. However, the COVID group demonstrated significantly higher fatigue levels compared to HC group (*T = 1093, p = 2*.*86e-07*). No significant difference was observed in any global morphometry metrics (*p > 0*.*05*). The central tendency measures for each measure along with statistical results are listed in Table 1 (see Supplementary Materials, Figure S1 for a visual assessment of group-wise distributions of GMV, WMV, CSFV and TIV).

The COVID survivors demonstrated significantly higher GMV compared to HCs. Figure 1 (left) shows two significant clusters exhibiting GMV differences between the groups in axial, sagittal and coronal planes. The clusters comprised of anatomical regions majorly from the *Limbic System* and *Basal Ganglia*. The multi-slice axial view (Figure 1, right) is provided with labels and arrows identifying some of these regions. More specifically, *bilateral hippocampus (Hc), left amygdala (Amg), insula (AIns, PIns), right entorhinal area (Ent*.) and *Parahippocampal gyrus (PHG)* from the *Limbic System* can be observed with higher GMV in the COVID group. From the *Basal Ganglia – left putamen* and *pallidum* also demonstrated higher GMV among the survivors compared to the HC group. The clusters survived a non-stationary cluster extent threshold of *k*_*E*_ = 1000 voxels with maximum peak voxel *t-*statistic values of *t*_*peak*_ *= 4*.*42* and *4*.*03*, respectively.

**Figure 1.**
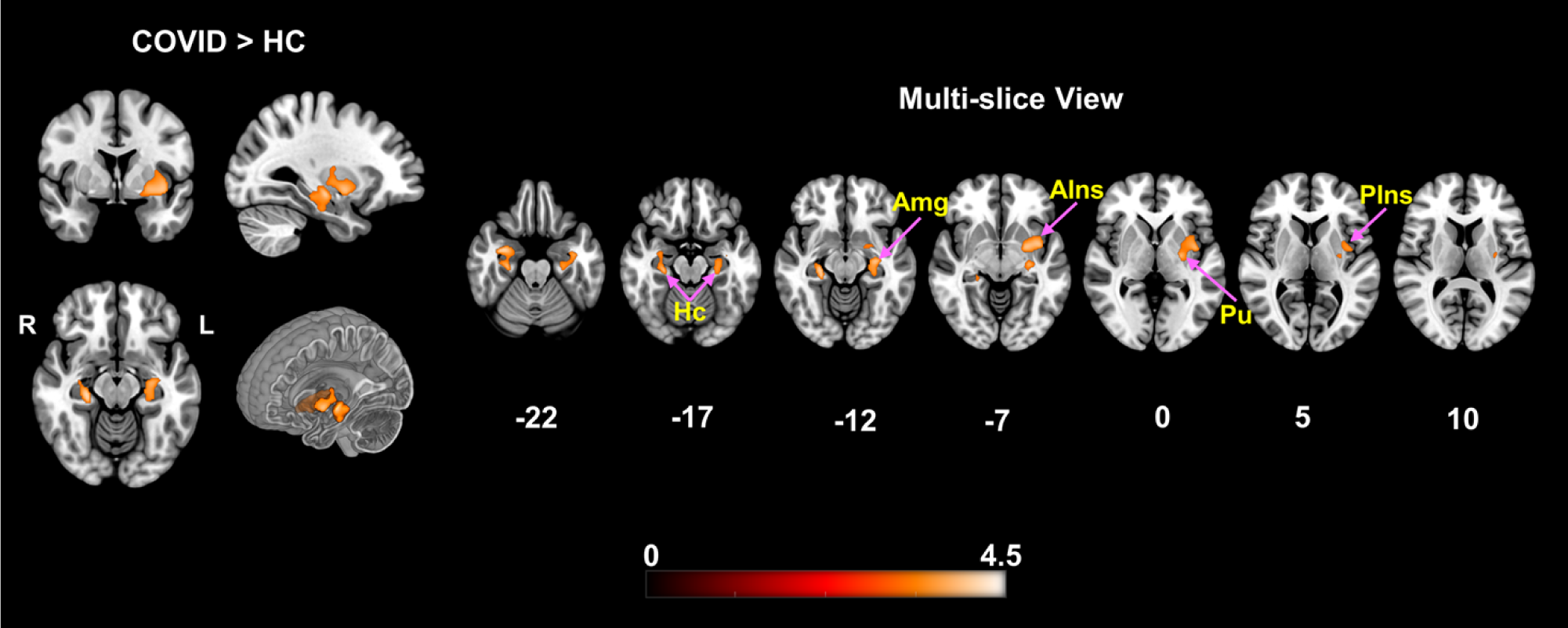
VBM demonstrating significantly higher gray matter volume in COVID-19 subjects compared to HC. Two significant clusters were observed, comprising of several deep brain structures: *Right* – *Hc, Am, Ent, PHG, VDC* and *Left – Hc, VDC, Pu, Pd, Am, PP, AIns, PIns*. The clusters surviving *FWE* correction, consisted of 1000 and 1968 voxels, with exact corrected p-values of *p = 0*.*017* and 0.023 at MNI coordinates: *[34 -4 -24]* and *[-28 -16 -10]*, respectively. The orthogonal slices on the left show the difference maps along with a cut-to-depth volume rendered image to better visualize the spatial extent of the anatomical locations comprising the cluster. The multi-slice image on the right shows finer slices (Z-slice gap >= 5) to highlight and assess the structural regions with significantly higher GMV. The cluster extends from inferior to superior Z-slices spanning from *Hc* to *AIns, Pu, and PIns* regions, respectively. Some of the relevant brain regions with significant differences have been pointed within the figure with purple arrows. The colorbar represents t-score values from the group level contrast. Keys: *Hc = Hippocampus, Am = Amygdala, Ent = Entorhinal Area, PHG = Parahippocampal Gyrus, VDC = Ventral Diencephalon, Pu = Putamen, Pd = Pallidum, PP = Planum Polare, AIns = Anterior Insula* and *PIns = Posterior Insula*.

For the correlation analysis using self-reported fatigue scores, the subset of participants (n_COVID_ = 33, n_HC_ = 18) demonstrated significantly positive correlation with GMV in *Posterior Cingulate Cortex (PCC), Precuneus (PRC)* and *Superior Parietal Lobule (SPL)*, particularly. The top left image in Figure 2 shows the significant cluster that comprised of these regions (see Figure 2 bottom row, for a multi-slice view). The scatter plot (Figure 2, top right) demonstrates the linear relationship *(Spearman’s ρ = 0*.*34, p = 0*.*016)* across the whole group between fatigue scores and the GMV of each subject within the cluster. The scatter plot combines data from both groups, and it can be noted that the light pink dots (COVID subjects) have a significantly higher effect compared to the cyan dots (HC subjects). Therefore, while GMV is positively correlated with fatigue across both groups, the overall trend is primarily driven by the effects from the COVID group.

**Figure 2.**
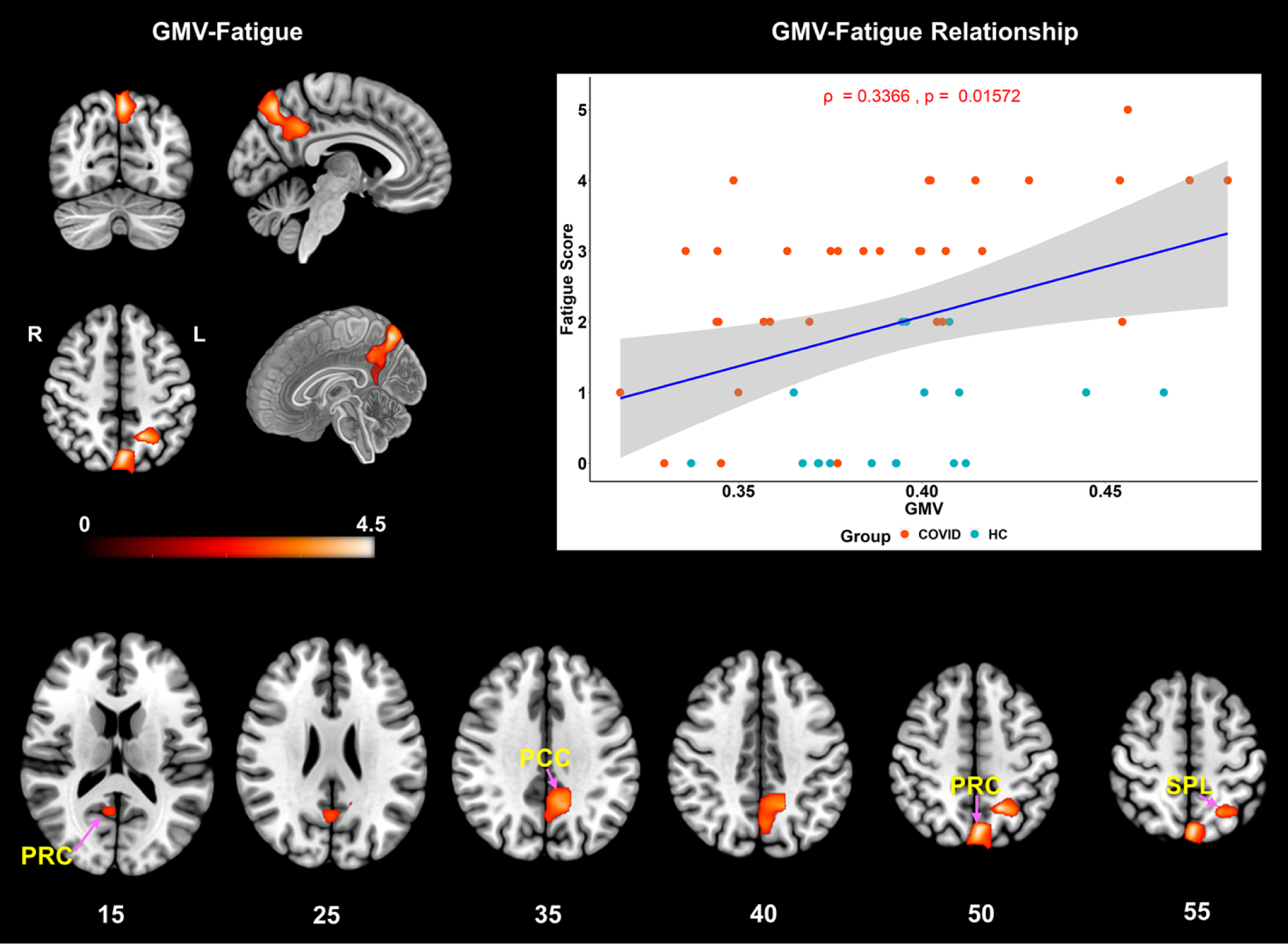
VBM demonstrating significantly positive correlation with fatigue scores across the whole group. The significant cluster (top-left) consisting of 3547 voxels (*k*_*E*_ *= 3547*), comprised of: *Left – PCC*, PRC, SPL and *Right - PRC* with peak t-score of *4*.*74* and exact corrected p-value of *p*_*FWE*_ = 0.019 at MNI coordinates: *[-16 -54 48]*. The axial slices (bottom) show the spatial extent of the same cluster over finer slices. The scatter plot (top-right) with the linear regression line shows significant positive correlation of GMV with self-reported fatigue score across the whole group *(ρ = 0*.*34, p = 0*.*016, r*^*2*^ *= 0*.*11)*. Please note, the *ρ* represents Spearman’s rank-order correlation coefficient. The light pink colored dots represent the COVID subjects, and the cyan dots represent the HCs. The COVID group clearly demonstrates higher effects than the HC group. Please note, the GMV in the x-axis represents the residuals plus the mean GMV of the cluster across subjects added back after linear regression. Keys: *PCC = Posterior Cingulate Cortex, PRC = Precuneus, SPL = Superior Parietal Lobule*. The linear plot (blue) represents the least squares regression line (best fit), and the shaded gray area represents the 95% confidence interval.

## Discussion

The VBM results support our hypothesis that COVID survivors, now PCR negative, demonstrate altered GMV compared to HCs even 2 weeks after hospital discharge. For the single case reports in the literature – hyperintensity in FLAIR images can arise from several sources including ischemia, micro-hemorrhages, and damage to vasculature, commonly observed in acute COVID patients and these neurological disturbances can modulate tissue volumes. We observed higher GMV in the *left Hc* and *Amg* at the group level, which aligns with a case report of hyperintensities in the *left Hc* and *Amg* from FLAIR images in an older patient (Male, 56 years) [2]. Recent neuroimaging studies have also reported higher GMV among COVID cohorts, in the *bilateral Hc* after 3 month-follow-up [11], as well as, in *bilateral Hc* and *Amg* in another follow-up study (3 and 6 months) [12]. Interestingly, Tu et al., 2021 also report significant correlation of the *left Hc* and *Amg* with PCL-5 scores, demonstrating stress related structural changes in these regions. On the contrary, cortical thickness was reported to be reduced in the *left Hc* after a 3 month-follow-up study [9] and in the *Amg* after infection with COVID-19 in a longitudinal study [10].

We also observed higher GMV in *AIns* and *PIns* regions indicating GMV alterations in *insular* lobes, which were reported to be hyperintense in specific patients [6]. Similarly, higher GMV was also observed at the group level in the *bilateral Ins* [11], while, on the other hand, others reported reduced cortical thickness in *insular* lobes [9, 10]. Early reports from severely acute hospitalized patients also mention hyperintense lesions in the *brain stem* and *basal ganglia* [7]. Our results also show higher GMV in *VDC, Pu*, and *Pd* which constitute major parts of the *basal ganglia* and *sub-cortical* system. Therefore, the group level estimates from our VBM analysis converge with acutely ill ‘individual’ COVID patient findings and maintain consistency with current neuroimaging reports from moderate to large samples of recovering COVID ‘groups’.

Although not statistically significant (*p > 0*.*05*), it is worth mentioning that the mean compartmental CSFV tended to be higher in the COVID group compared to the HCs (see *Table 1* and Figure S1 from Supplementary Materials). This seems to align with the recent UK-Biobank longitudinal study (n = 401), who reported increased CSFV after COVID infection [10]. Our data indicates more variation in brain volumes among the COVID patients, with higher standard deviation (SD) in GMV, WMV and TIV (see *Table 1*). This might arise from varying levels of tissue swelling due to acute infection among surviving patients. However, the underlying neurophysiology that elicit such changes is still unclear. Highly prevalent acute stage neurological damage from CNS viral or vascular pathologies can cause local changes in tissue concentration. Transient reduction in cerebral blood flow (CBF) can also cause gray matter concentration to increase due to change in hydration levels[23]. Overall, continued brain swelling from neuro-vascular injuries may explain why we observed locally higher GMV in the COVID group even 2 weeks after testing negative.

Our results also support the hypothesis that fatigue levels experienced by the COVID survivors will be higher compared to the HC group and GMV will correlate more strongly with self-reported fatigue in this group. A rising concern with recovering patients has been the manifestation of a sequelae of symptoms that persist several months along the recovery timeline [13-15]. Fatigue was the highest reported symptom in these studies among others, including lack of attention, delayed recovery of loss of sense of smell and taste. We asked the healthy subjects and surviving patients, what level of fatigue is disrupting their daily work? Based on their reported score, we observed significantly higher levels of fatigue within the COVID group when compared to HCs. We also observe from Figure 2 that GMV in *PCC, PRC* and *SPL* regions are more strongly correlated to fatigue in COVID survivors compared to healthy controls. This could also indicate a link to high percentage of survivors experiencing fatigue during and post recovery, eventually leading to functional or cognitive disruption. This is also typical of neurodegenerative populations and these regions have been found to be related to fatigue. For example, higher metabolic activity within *PCC* has been shown to be positively correlated with higher fatigue levels in Parkinson’s patients [24]. *PCC* and *SPL* have also been shown to be associated with fatigue in patients with chronic fatigue syndrome (CFS) [25, 26]. Moreover, atrophy in the *parietal* lobe has been shown to be associated with fatigue among multiple sclerosis (MS) patients [27, 28].

Not shown here is another cluster (did not survive non-stationary clustering threshold, please see Supplementary Material, Figure S2) that consisted of brain regions from the *cholinergic output (BsF, AcA)* from the *ventral BG* and *orbitofrontal cortex (MOG, GrE), ACG* and *MFC* from the *ventromedial prefrontal cortex (vmPFC)*. When the cluster GMV was linearly regressed against fatigue, a significant positive correlation (*ρ = 0*.*41, p = 0*.*0028*) was observed (see Supplementary Material, Figure S2). Interestingly, the *BG* and *vmPFC* regions have also been shown to be functionally associated with fatigue [29-34]. We observed this effect with 18 HCs and 33 COVID subjects and our expectation is that with a bigger sample size in both groups, this cluster would survive and therefore it may be of relevance to fatigue related effects among survivors.

In conclusion, our results highlight group level effects in surviving COVID-negative patients that match with single patient case studies, as well as several neuroimaging studies from surviving COVID-19 cohorts. We have shown significant GMV alterations in multiple brain regions from the *limbic system* and *basal ganglia* and further showed positive associations of regional volume with self-reported fatigue at work. More importantly, these regions can also be modulated by neuronal damage and characterized from functional neuroimaging relating to fatigue, pain, emotion, attention, and somatosensory processing [35-38].

## Limitations and Future Directions

Despite our emphasis on group level analysis, we understand that in a clinical setting, it may have little transferability, owing to idiosyncrasies associated with each patient. A possible approach to address this issue would be to compare patient specific VBM against a sufficiently sized control sub-group randomly selected from a larger cohort [39]. However, it may not be very practical in a clinical setting unless a well-designed control dataset is available for a clinician. Nevertheless, our group level results from a single site seem to match single patient findings quite well, especially when they were collected from several centers across different countries [40]. Moreover, currently, we only have 46 COVID subjects. While we do observe significant effects, we still need a larger sample size to verify the main effects more conclusively. Another concern we have is the reversibility or transient nature of some effects. These patients were scanned two weeks after hospital discharge. It is possible some critical effects may have already disappeared through recovery or reversible neurological processes. Therefore, a better approach could be to scan the patients at onset, during and after recovery along with behavioral parameters to assess any possible trends unique to the neurological pathology in COVID patients.

## Data Availability

All data produced in the present study are available upon reasonable request to the authors

## CRediT author Statement

**Rakibul Hafiz**: Methodology, Software, Formal Analysis, Data Curation, Writing – Original Draft, Review and Editing.

**Tapan K. Gandhi:** Conceptualization, Investigation, Resources, Supervision, Writing – Review and Editing.

**Sapna Mishra:** Investigation, Resources, Data Curation, Writing – Review and Editing.

**Alok Prasad:** Writing – Review and Editing.

**Vidur Mahajan:** Writing – Review and Editing.

**Xin Di:** Methodology, Writing – Review and Editing.

**Benjamin H. Natelson:** Writing – Review and Editing.

**Bharat Biswal:** Conceptualization, Resources, Project Administration, Supervision, Writing – Review and Editing.

## Acknowledgements

This study was supported by an NIH grant (R01AT009829).

## Notes

### Competing Interest Statement

The authors have declared no competing interest.

### Funding Statement

This study was funded by an NIH grant (R01AT009829).

### Author Declarations

Ethics Committee/IRB of Indian Institute of Technology (IIT), Delhi, India, gave ethical approval for this work.

### Summary of Updates

The revised version of the manuscript has undergone the following changes: 1. Abstract has been rewritten in a structured format. 2. Introduction has been made more concise. 3. Additional clinical information pertaining to COVID survivors has been added under Participants in Materials and Methods. 4. Non-stationary cluster-based thresholding has been adopted at a more conservative height threshold of p < 0.001 for the t-test 5. Fatigue scores from Healthy group has also been added and group level difference with COVID group has been reported. 6. The voxelwise correlation analysis has been updated with scores from both Healthy and COVID groups and new results have been reported. 7. Discussion on the cluster reported in the previous manuscript has been added in the Discussion section and relevant results added in the Supplementary Material document as Figure S2 8. Figures 1 and 2 have been combined to make Figure 1 in the revised manuscript. 9. Figure 3 in the previous manuscript has been moved to the Supplementary Material document as Figure S1

