## Supplementary Material for "Higher Limbic and Basal Ganglia volumes in surviving COVID-negative patients and the relations to fatigue"

**Supplementary Materials**

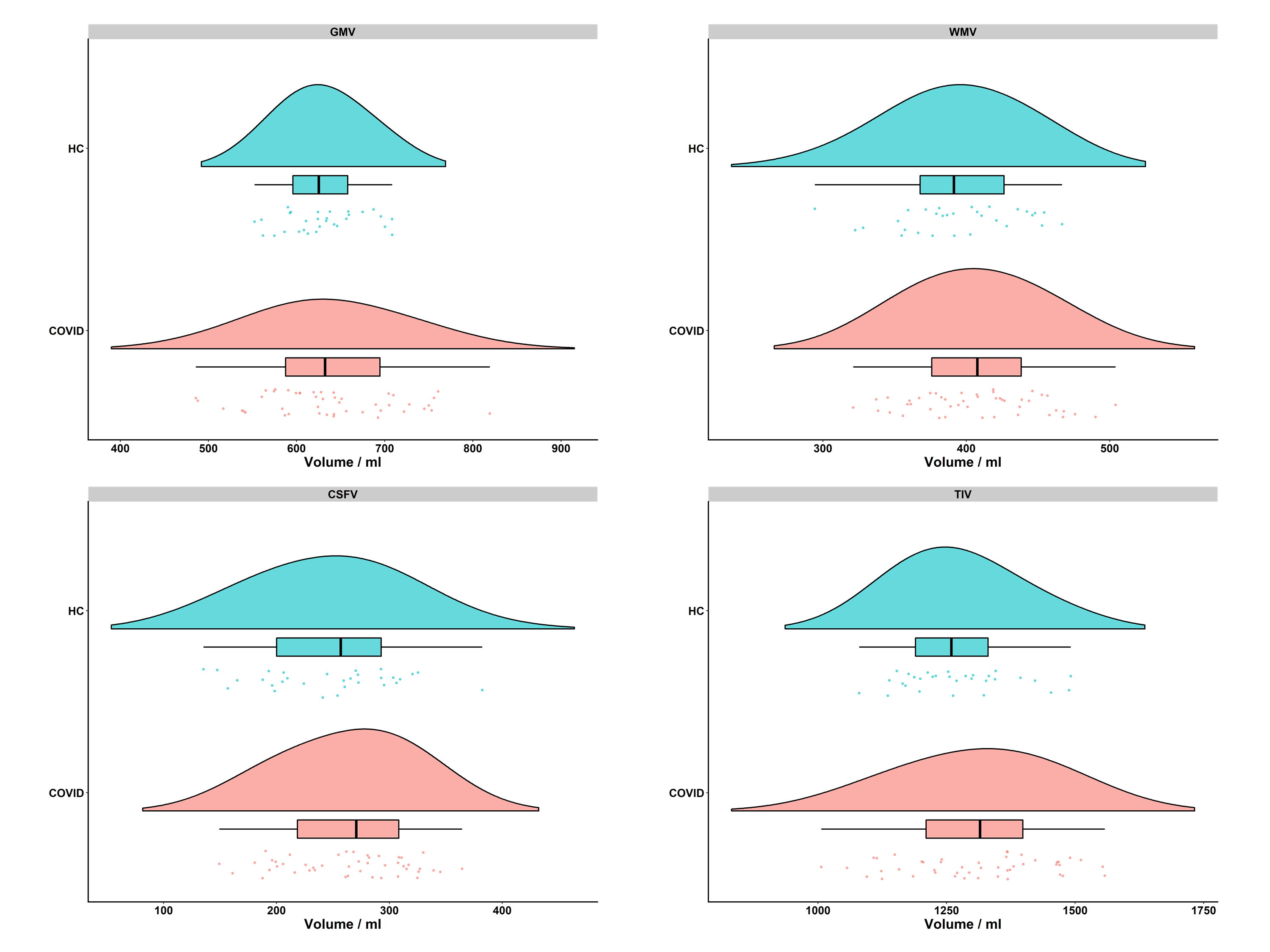

**Figure S1.** **‘Raincloud’ plots of compartmental and total intracranial volume distribution and central tendency measures from each group.** The figure shows the flat violin, box and dot plots all within the same frame for each group to assess the central tendency measures, such as the mean, standard deviation and inter-quartile range of compartmental volumes (top row and bottom left), as well as the TIV (bottom right) in each group. The cyan color represents the HC group, and the light pink color represents the COVID group in each plot. The box plot shows the tendency of higher group averaged WMV, CSFV and TIV in the COVID group and marginally lower GMV compared to the HC group. The individual volume values that drive this distribution can be assessed from the dot plot and the slight shift of the light-pink violin distribution plot to the right. GMV = Gray Matter Volume, WMV = White Matter Volume, CSFV = Cerebrospinal Fluid Volume, TIV = Total Intracranial Volume

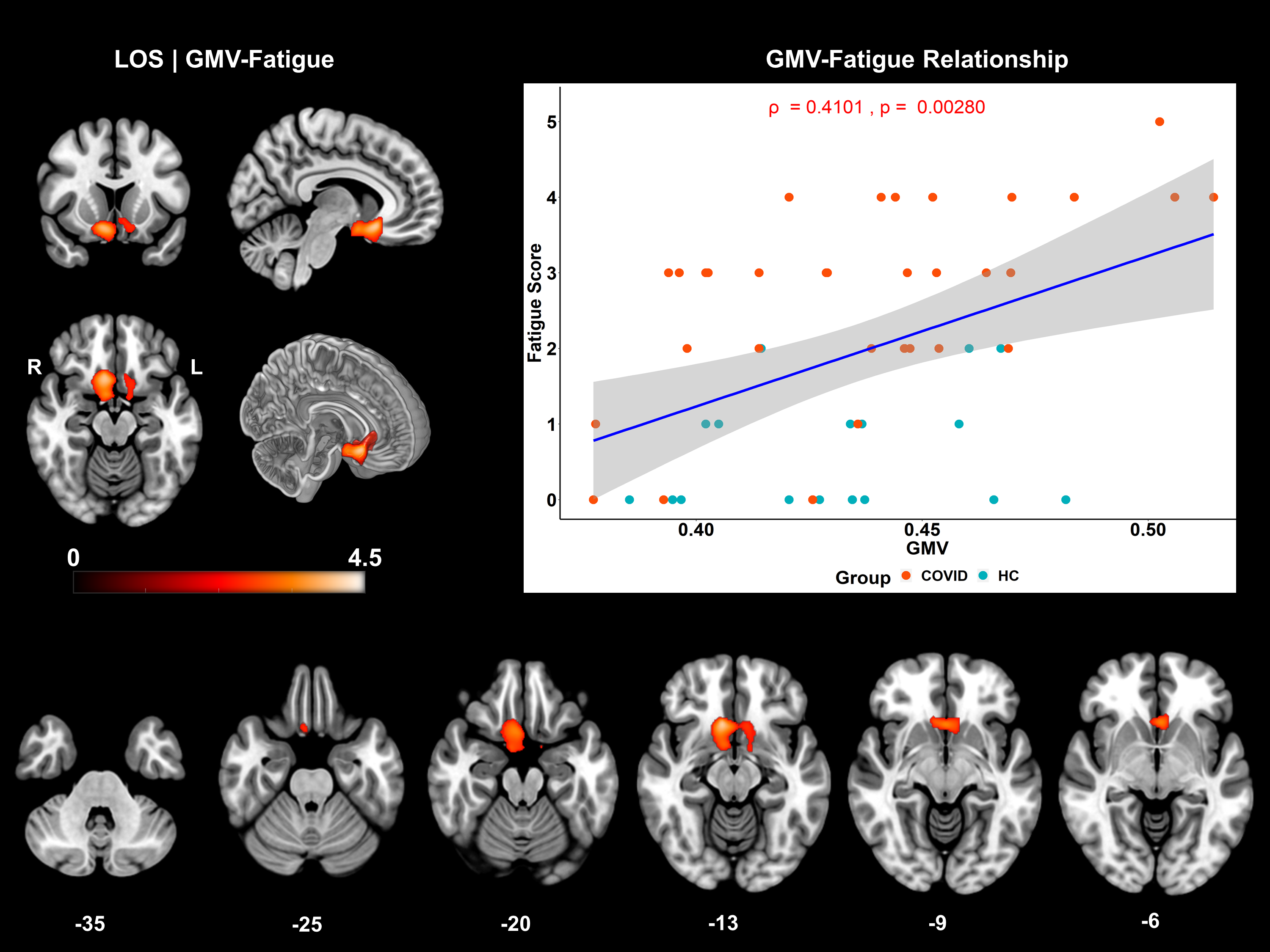

**Figure S2.** **VBM demonstrating positive correlation with fatigue scores across the whole group.** The cluster (top-left) (1796 voxels) did not survive the non-stationary cluster threshold (k_E_ = 3547 voxels). However, when the residual plus mean GMV across group was regressed against fatigue scores, a significant effect was observed (top-right). The cluster comprises of *bilateral – Subcallosal Area (ScA), Accumbens Area (AcA), Mid-orbital Gyrus (MOG), Anterior Cingulate Gyrus (ACG), Medial Frontal Cortex (MFC), Gyrus Rectus (GRe), Caudate (Cd), Putamen (Pu) and Bilateral – Ventral Diencephalon (VDC) and Right – Basal Forebrain (BsF), Amygdala (Amg), Entorhinal Area (EnA) and Parahippocampal Gyrus (PHG)*. A multi-slice axial view has also been added to showcase the spatial extent of this cluster. The linear plot (blue) represents the least squares regression line (best fit), and the shaded gray area represents the 95% confidence interval.
